# Knowledge, attitude and practice of primary school teachers toward epilepsy in Khartoum city, Sudan: A cross-sectional Study

**DOI:** 10.1101/2022.10.10.22280849

**Authors:** Abubaker A MohamedSharif, Isra Bdraldein Salih Mohammed, Abubaker E.A. Koko

## Abstract

**Objectives:** The aim is to assess the knowledge, attitude and practice of the primary school teachers toward.

**Methods:** A descriptive cross-sectional study was conducted among primary school teachers in Khartoum using self-administered, closed ended questionnaires.

**Results:** Almost all teachers have heard about epilepsy before. Most of them believed that epilepsy was due to genetic factors (25.4%). (74.6%) of the participants thought that epilepsy was a contagious disease. Concerning practice, (26.5%) of the teachers who did first aid, they carried out all the steps of management truly.

**Conclusion:** Teachers generally have some knowledge about epilepsy, but there were also deficiencies in their management measures. Most of them heard about epilepsy from community reflecting the need for training sessions and a national program to raise their awareness and attitude.

## Background

Epilepsy is considered one of the most important chronic neurological disorders (1,2). It is characterized by synchronous recurrent unprovoked seizures due to uncontrolled electrical discharges from the brain neurons (2, 3).

The disease has high prevalence as it is estimated that about 50 million people are affected worldwide (8). The global burden of the disease is about 1% in both industrialized and developing countries, being more in the developing one such as Sudan (80-85% of the cases) (1,2), with high incidence 100-190 per 100,000 people per year in developing countries (5).

Epilepsy affects lifestyle of primary school epileptic pupils (9,10,11) both academically because of social barriers (9) and psychologically including anxiety, depression and social stigma (12,11,6,13,14,15). In addition, epilepsy has been associated with low self-esteem (13) and educational achievement (16,17,18,19). Furthermore, negative attitude of non-epileptic children and primary school teachers toward epilepsy has been associated with a negative impact on children with epilepsy. (1,9,20,21).

Social stigma of epilepsy can be very difficult to overcome causing many of patients avoid seeking medical advice and increasing the “treatment gap” which exaggerates the condition more than the convulsions and the disease itself (22). Disappointingly stigmatization and social discrimination applied by the community will increase the burden of this disease regarding management and other aspects of disease (6,13).

Community involvement, especially of primary school teachers must play a great role in eradication of this social dis acceptance, because they are regarded one of the educated sectors in the community (16,25,26,27) and by their fundamental participation in the education of the epileptic pupils (19). We need to minimize epilepsy-associated social stigma (16) by good lightening of the main problem which is teacher’s awareness about epilepsy causes, management, and attitude (16,25).

Understanding the misconceptions and false beliefs of teachers about epilepsy is a vital point, putting in mind that there are no formal instructions about epilepsy during their training (1,28). Several studies have shown that despite improvement in perception (29,30) and community attitude, there are still misconceptions about epilepsy (12,31). So there is need to design programs to antagonize these misunderstandings.

Up to date, there is still a little researches had been conducted in this part of the disease especially in Middle East (6,32,33) and Sudan (1).

## Objectives

This study aimed to investigate and examine the knowledge, attitude, and practice of primary school teachers toward epilepsy regarding causes, clinical presentations and management of convulsing epileptic pupils.

## Methods

### Study design and population

A descriptive cross-sectional study was conducted on January 2018. All teachers at pensionable age employed in or training at registered public schools and present at the time of the study were eligible to participate in the study. We exclude those who refused to participate and did not give verbal consent.

### Study area

Khartoum City, consist of 18 districts with 58 public schools, 82 private schools and two universities.

### Sampling (size and technique)

The sample size was calculated using the equation n = [DEFF*Np(1-p)]/ [(d2/Z21-α/2*(N-1)+p*(1-p)] where n = sample size, z = level of confidence = 1.96 (95% CI), P = prevalence., we assumed 50% prevalence of inadequate knowledge, attitude and practice. Therefore, P = 0.5, q = 1–p = 0.5, d = desired margin of error = 0.05. The sample size was found to be 384 and 33 teachers refused to participate in the study so n=350.

Questionnaires were given to the directors of schools to distribute them to all eligible teachers to complete them and then returned to research assistant. Instructions on how to complete questionnaires with few explanations were given avoiding leads. Study was carried out in 18 schools as clusters which were selected randomly (the sampling technique used was cluster random sampling), then a total coverage to the selected schools was done. Each school involved about 20 teachers and total sample of 350 was taken.

### Data collection

The data were collected using structured closed ended self-administered questionnaires and the questions were designed to cover KAP with respect to epilepsy. To maintain the standard of the survey material other similar studies were identified and relevant questions were selected and modified to make them appropriate to the local culture. The selected questions were first forward translated into Arabic the official language of the country, and then back translated into the English using the standard translation procedure. The translated questionnaire was then pretested randomly on selected school teachers which had helped to further redirect and rephrase some questions. Participants were asked to complete a self-administered questionnaire, consisting of 21 questions. Consisting of four parts, sociodemographic data, knowledge questions, attitudes and practices. Each questionnaire was checked for completion, assigned a code, kept in a secured file and entered into an electronic database for analysis. Data was analyzed using Statistical Package of the Social Sciences (SPSS) software v21.0 (IBM SPSS Statistics). Frequencies were calculated for all independent (age, gender, educational level, experiences) and dependent variables.

### Ethical consideration

Ethics approval and consent to participate:

The study was approved by the Institutional Review Board of the Faculty of Medicine of University of Khartoum in accordance with the Declaration of Helsinki.

To ensure adherence to ethical guidelines, several measures were adopted while conducting this study;

- Study participants were recruited from the community (in-person and online), and no incentives were offered to the participants in return for their participation.
- Informed consent was provided to the participants and obtained before filling the survey.
- Participants were informed that their participation in this study is voluntary, no incentives or compensations will be offered in return, and that they have the right to withdraw from the study at any stage. The scientific value of their participation was explained in the informed consent.
- The contact information of the principal investigators was provided for participants.

All the participants’ information was kept private by keeping it in a secured folder in a password-protected computer owned by the study investigators. No information was shared with any other individuals or entities.

## Results

The study was conducted in (350) primary school teachers with response rate of 91.1%. The mean age of them was found to be (47) years old with an interval of (20-70) years with females being the predominant sex (Male: female ratio was 1:2.9). Most of them were married (68.9%) and reported that they were university graduate (60.3%) and. Thirty teachers (22.8%) had teaching experience of (10-20) years (see table 1).

**Table 1:**
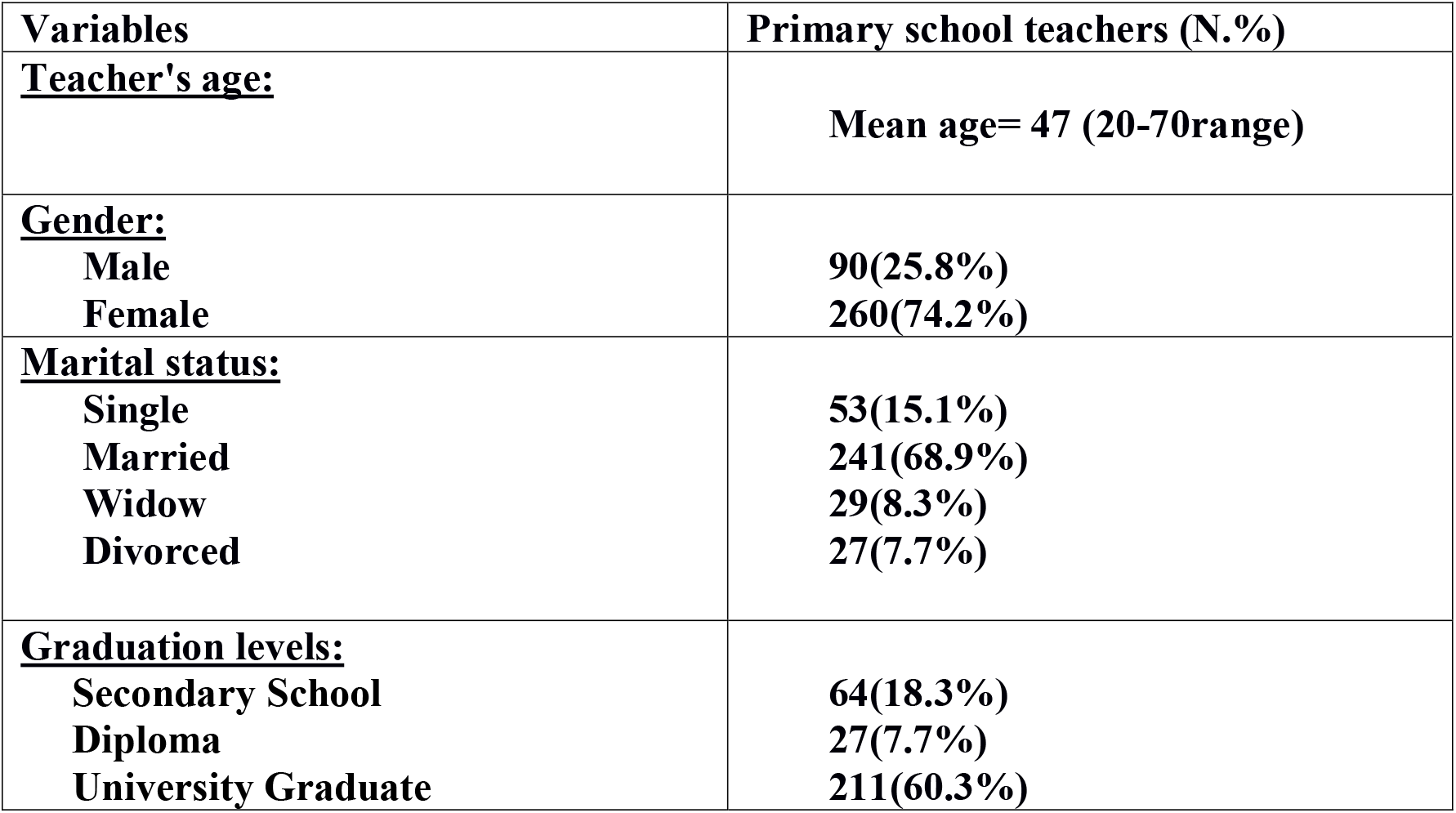
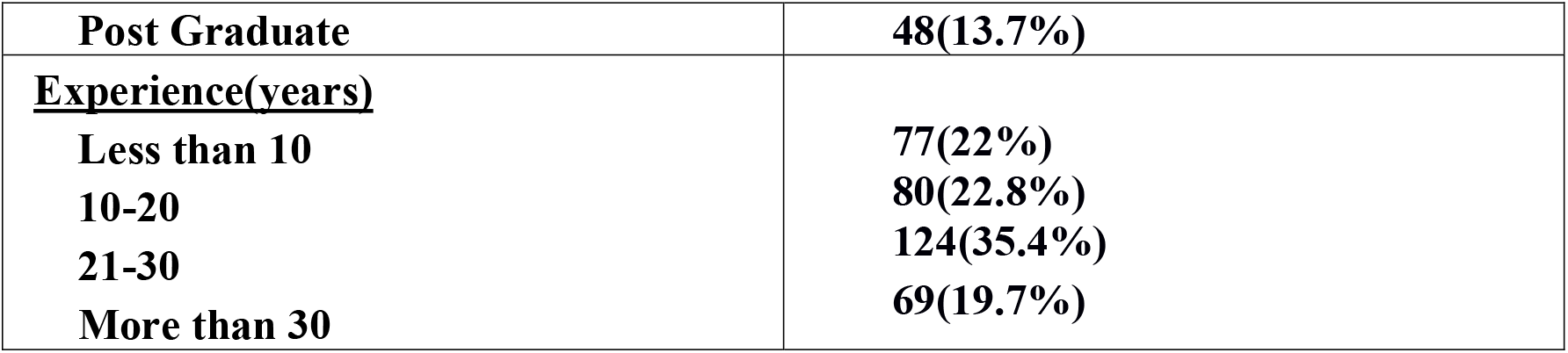
(Primary school teacher’s Sociodemographic data)

### Knowledge about epilepsy

As all the participants heard about epilepsy before, they reported that community (32.1%) and media (29.1%) were the main source of information and few of them (7.2%) gained their information about epilepsy from Health Care Givers. Regarding the causes of epilepsy, most of the participants believed that epilepsy was due to genetic factors 88(25.4%) and head injuries 63(18.2%), while 44(12.7%) of them thought it was due to evil spirits. About 182(51.9%) of the participants thought that the most useful treatment was medical treatment in form of tabs and injections and minority of them; 24(6.9%) thought of traditional treatment and 21(6.1%) believed of electrical shock to be the best one (see table 2).

**Table 2:** (primary school teacher’s knowledge about causes and treatment of epilepsy)

| **Variables** | **Primary school teachers (N.%)** |
| --- | --- |
| **<u>Causes of epilepsy:</u>** |  |
| **Genetic Factors** | **88(25.4%)** |
| **Head Injuries** | **63(18.2%)** |
| **Malaria and fever's causes** | **25(7.2%)** |
| **Evil spirits** | **44(12.7%)** |
| **Inflammation of the brain tissue and meningitis** | **59(17.0%)** |
| **Brain Tumors** | **24(6.9%)** |
| **Increased electrical activity of the brain** | **37(10.7%)** |
| **I Don't Know** | **6(1.7%)** |
| **<u>Treatment of epilepsy:</u>** |  |
| **Medical Treatment ( Tabs Or Injections )** | **182(51.9%)** |
| **Surgical Intervention** | **35(9.9%)** |
| **Electrical Shocks** | **21(6.1%)** |
| **Traditional Treatment** | **24(6.9%)** |
| **I Don't Know** | **77(22.1%)** |

261(74.6%) of the participants they considered epilepsy as a contagious disease. Majority of the participants thought that convulsions should have to occur during an episode of epilepsy (65.1%) and about 82 teachers (23.5%) thought that it wasn’t necessarily to occur. 165(47.2%) of them believed that epileptic pupils can participate in all sport activities as healthy one can do while 118(33.7%) thought that they can’t (see table 3).

**Table 3:** (Primary school teacher’s knowledge)

| **Variables** | **Primary school teachers(N.%)** |
| --- | --- |
| **<u>Convulsion should have to occur during episode of epilepsy</u>**<br>Yes<br>No<br>I don't know | 228(65.1%)<br>82(23.5%)<br>40(11.4%) |
| **<u>Think epilepsy is a contagious disease:</u>**<br>Yes<br>No<br>I don't know | 41(11.7%)<br>261(74.6%)<br>48(13.7%) |
| **<u>Epileptics can contribute in all sports activities:</u>**<br>Yes<br>No<br>I don't know | 165(47.2%)<br>118(33.7%)<br>67(19.1%) |

### Attitude towards epilepsy and epileptic pupils

Concerning attitude of the participants, most of them (77.1%) allow epileptic patients to learn with other healthy one. Those whom don’t allow, more than (51.5%) of them they thought that epileptic patients can harm others (12.1%) had difficulties to learn (12.1%) not qualified to work later on and (21.2%) they thought that others can harm them. Most of them they thought that epileptic patients were not socially separated by their colleges (71.7%). Almost all the participant 328(93.7%) they show empathy to a convulsing pupil and a very minor group 19(5.6%) feel afraid of him. 255(73%) of teachers revised for an epileptic pupil who missed a class because of his illness (see table 4).

**Table 4:** (Primary school teacher’s attitude toward epileptic pupils)

| **Variables** | **Primary school teachers ((N.%)** |
| --- | --- |
| **<u>Allowing epileptic pupils to learn with other healthy one</u>**<br>Yes<br>No<br>I don't know | 270(77.1%)<br>78(22.3%)<br>2(0.6%) |
| **<u>Reasons to disallow them to learn:</u>**<br>Can Harm Others<br>Has Difficulties To Learn<br>Are Not Qualified To Work<br>Other Can Harm And Cause Problems To Him | 40(51.5%)<br>9(12.1%)<br>9(12.1%)<br>17(21.2%) |
| **<u>Feeling of teachers toward epileptic pupils:</u>**<br>Fear | 19(5.6%) |
| **Empathy** | **328(93.7%)** |
| **Ignorance** | **3(0.8%)** |
| **<u>Social separation by their colleges:</u>** |  |
| **Yes** | **69(19.7%)** |
| **No** | **251(71.7%)** |
| **I don't know** | **30(8.6%)** |
| **<u>Revise for epileptic pupil who missed a class because of his illness:</u>** |  |
| **Yes** | **255(73%)** |
| **No** | **95(27%)** |

### Practice of teachers regarding epilepsy

110(31.5%) of the teachers taught an epileptic pupil before. About 64(18.2%) of the teachers did first aid management to an epileptic patient and from this percent 64(18.2%); 22(34.4%) remove the sharp object and secure the patient, 9(14%) calculate the duration of convulsion, 14(21.9%) put the patient in his lateral side, 19(29.7%) ensure airway patency and 17(26.5%) did all the management steps. Regarding those who didn’t carry out first aid before have knowledge of following: 104(29.7%) removing the sharp objects and secure the patient 70(20%) putting a piece of clothes or spoon in his mouth (see table 5).

**Table 5:** (practice of the primary school teachers)

| **Variables** | **Primary school teachers (N.%)** |
| --- | --- |
| **<u>Taught an epileptic pupil before:</u>** |  |
| **Yes** | **110(31.5%)** |
| **No** | **240(68.5%)** |
| **<u>Did first aid during convulsion before:</u>** |  |
| **Yes** | **64(18.2%)** |
| **No** | **286(81.8%)** |
| **<u>Management steps teachers did before:</u>** |  |
| **Removing the sharp objects and secure the patient** | **22(34.4%)** |
| **Calculate the time of convulsion** | **9(14%)** |
| **Putting the patient on his lateral side** | **14(21.9%)** |
| **Ensuring airway patency and giving first aid management** | **19(29.7%)** |
| **All steps together** | **17(26.5%)** |
| **<u>Management steps teachers should do:</u>** |  |
| **Removing the sharp objects and secure the patient** | **104(29.7%)** |
| **I'll run away and don't intervene** | **15(4.3%)** |
| **Putting a piece of clothes or spoon in his mouth** | **70(20.0%)** |
| **Calculate the time of convulsion** | **24(6.9%)** |
| **Putting the patient on his lateral side** | **27(7.7%)** |
| **Calling the ambulance without intervening** | **29(8.3%)** |
| **Ensuring airway patency and giving first aid management** | **50(14.3%)** |
| **Control and stop convulsions by tie him with rob** | **10(2.9%)** |
| **I don't know** | **18(5.1%)** |

## Discussion

This research was done to assess the degree of knowledge, attitude and practice of primary school teachers toward epileptic pupils to design a program for raising their awareness about this disease because no formal instructions were given to them during their training

Majority of teachers in our study got their information about epilepsy from the community 32.1% and media 29.1% highlighting the necessity to encourage health educational public campaign to be community based and encourage media to disseminate knowledge about the disease. In addition, this was considered to be a good thing, as media became a popular way nowadays and almost all people have easy access to media. Our result was similar to that obtained from India and Thailand (39,41,11) and in contrast to study done in Ethiopia 2016 in which public media played a minor role (39). Few of our participants gained their information about epilepsy for health care workers 7.2%, this in contrast to that done in Ethiopian teachers (39), in which health care workers represent a common source of information.

Some previous studies reported that teachers had high knowledge about epilepsy (38,42). Despite that, they had a defected knowledge and some misconceptions about the disease (35,38,43,44). One of this misinformation was the believed that epilepsy was a contagious disease 74% (19,43,41,45,46, 47,48,48,50) and in contrast to that done in Saudi Arabia (47), this thought plays a miserable role in increase the stigma and social barriers and this should be a warning alarm to establish an interventional program to the raise teacher’s awareness. This result was similar to other studies (43,51).

On other hand, a significant percentage (28.2%, 44%) of teachers participating in previous studies (1,38) reported that they believed that epilepsy affect growth and development of children, despite their greater years of experience. This misconception also increases stigmatization and discrimination more and more. The importance of teacher’s attitude is that it affects other children attitude affecting their own development.

Other aspect defected in their knowledge, evil spirits being one of the causes of epilepsy and this also contribute in increasing social stigma (19). Few of participants 12.7% still believe in evil spirits as a cause of epilepsy compared to 21.1% reported in Sudan 2017 (59), 26% reported in Nigerian teachers (40).

Regarding causes of epilepsy, a part of those didn’t know the cause, most of our participants, report genetics 25.4% and head injuries 18.2% as a common cause of epilepsy compared to study done in Sudan 2017 in which 78% of teachers believe epilepsy was due to neurological problems and slightly less than this value reported in Ethiopia (39) and Pakistan (37).

More than half of our teachers thought of medical treatment to be the most useful one. This proportion is lower than that reported in Nigerian which was found to be two-third (62). In Ethiopia 98% of participants thought about medical treatment as the most useful one. 6.9% of our participants believed of traditional one as best in contrast to study done in Zambia reported that about 84.5% of teachers thought of traditional healers as the best rather than doctors (63) and also in contrast to another study done in Ethiopia which reported that near half of the teachers believed in those healers (39). In study done in Sudan, 9.4% believed in traditional treatment as a curative one compared to 51% who didn’t.

More than two-third of the teachers reported in our study that they allowed epileptic pupils to learn with their healthy colleges in the same classroom. Despite this high proportion, a significant percent of them (22.3%) didn’t allow that comparable to study done in Nigeria (7%), India (20.8%) and in Thailand (15.1%) (19,41,52). Even severely, there was study reported that 4.5% of teachers refused to teach epileptics at all (38) and another one reported 13% refused also (64). Our teachers whom didn’t allow epileptic pupils to learn with other healthy one, 51.5% of them believed that epileptics can harm other similar to study done in Egypt 46.6% (36) or might be not able to worker later on 12.1% similar to studies which reported that teachers thought epileptics can’t be a good teacher in the future and can’t succeed in their life (43,11,38). This believed reflect the great gap that teachers think of epilepsy as a disease affecting the cognitive functions of the patient as reported in similar study leading us to think of their inferior misconception about epilepsy (43).

Most of our participants 71.7% think epileptics were not socially separated by their colleges reflecting teachers with some positive attitude.

In our study, almost half of the teachers allow epileptics to participate and contribute in activities with healthy one similar to previous studies with high percentage also (38,63). Despite that we need to tell them that there must be some special limitations in those activities like swimming or driving.

Luckily, only 5.6% of teachers in this research felt afraid of epileptics when seeing them convulsing which is much lower than that reported in study done in Saudi Arabia (47) and more than ninety percent of teachers said that they felt sympathize with a fitting child, which is higher than that reported in Egypt 76.8%, Nigeria which reported that more than half of them afraid of those students, 32% of teachers afraid of epileptics in study done in India and 9.8% afraid of epileptics in study done in Thailand (19,41,52) reflecting good attitude of our teachers in this regard. The feeling of fear toward epileptics affects them both psycho socially and academically raising the stigmatization and discrimination upon them.

One-fourth of the teachers who did first aid before, carried out all the steps of management correctly. Unfortunately, some of them did the procedures hastily as reported in similar studies by pulling the tongue or putting objects like spoon inside the mouth (43,11,19,65,31, 38). Because of that, there must be school health services as the teachers being always in contact with epileptic pupils. Regarding these services, physicians also should contribute by sharing the knowledge and the medications that can help the teachers to deal with such a situation.

## Conclusion

This study assesses knowledge, attitude and practice among primary school teachers in Khartoum. In conclusion, the knowledge of the teachers varies at time of this research. There were misunderstandings and misconceptions in some aspects of the disease like causes of epilepsy being evil spirit and being a contagious disease and good knowledge reported in area of treatment. The more educated teachers about epilepsy the less likely for having negative attitude. Teachers of our study were considered to have negative attitude toward epilepsy which badly affects their awareness. Lack of awareness is a warning alarm of establishing educational programs about epilepsy. Bad attitude increases stigma and social barriers. The practice of teachers was considered to be in need of improvement as there are no formal instructions about epilepsy during their training.

## Data Availability

All data produced in the present study are available upon reasonable request to the authors

## Recommendations

We need further researches in context of knowledge of primary school teachers about epilepsy to explore areas of deficiencies that teachers suffered from. Also, we need these researches aiming to investigate about the practical aspect of epilepsy as teachers form the cornerstones in the educational process of those epileptic pupils. The need for educational health public programs is of the greatest importance recommended erasing all myths and misconceptions about epilepsy which is the main cause of the social stigmatization and discrimination.

## Declarations

## Acknowledgments

The authors would like to thank the faculty and staff at the Faculty of Medicine – University of Khartoum for their continuous support.

## Ethics approval

This study was approved by University of Khartoum, Faculty of medicine, community department.

To ensure adherence to ethical guidelines, several measures were adopted while conducting this study:

1. No incentives were offered to the participants in return for their participation.
2. Verbal consent was obtained from the participants before filling the questionnaire.
3. Participants were informed that their participation in this study is voluntary, no incentives or compensations will be offered in return, and that they have the right to withdraw from the study at any stage. The scientific value of their participation was explained in the verbal consent.
4. The contact information of the principal investigators was provided for participants.
5. All the participants’ information was kept private by keeping it in a secured folder in a password-protected computer owned by the study investigators. No information was shared with any other individuals or entities.

## Availability of data and materials

The datasets used and/or analyzed during the current study are available from the corresponding author on reasonable request.

## Authors’ contributions

AM: conception of this study. IM designed the study and formulated its methodology. AK and AM analyzed the data. AM and IM interpreted the data. IM contributed to the results section. IM and AM contributed to the discussion section. AK drafted the work and substantially revised it.

AK critically reviewed and approved the manuscript.

All authors listed have agreed to both be personally accountable for the author’s own contributions and to ensure that questions related to the accuracy or integrity of any part of the work, even ones in which the author was not personally involved, are appropriately investigated, resolved, and the resolution documented in the literature.

All authors listed read and approved the final manuscript.

